# The role of rare copy number variants in early-onset depression

**DOI:** 10.1101/2025.05.14.25327601

**Authors:** Charlotte A Dennison, Ida Sønderby, Miguel Garcia-Argibay, Victoria Powell, Amy Shakeshaft, Lucy Riglin, Elliott Rees, Michael O’Donovan, Henrik Larsson, Ole Andreassen, Alexandra Havdahl, Joanna Martin, Anita Thapar

## Abstract

**Background:** Depression is a highly heterogeneous condition. Depression with an onset in childhood and early adolescence has a worse clinical course, is more heritable, and shows a lower genetic correlation with other depression subtypes, than does later-onset depression. It is also more strongly associated with neurodevelopmental comorbidities and genetic liability to ADHD. Thus, we hypothesised that early-onset depression represents a distinctive “neurodevelopmental” depression subtype associated with an increased burden of rare copy number variants (CNVs) that are enriched in neurodevelopmental conditions. We tested this hypothesis using four population cohorts across the UK, Norway, and Sweden.

**Methods:** Participants were ascertained from four population cohorts across the UK, Norway, and Sweden. Early-onset depression was defined as a score >11 on the self-reported Short Mood and Feelings Questionnaire (SMFQ) between ages 10 and 14 years (cases n=5,994 vs controls n=26,388) and, for secondary analyses, using an ICD-10 criteria for major depressive disorder (MDD) with onset ≤14 years (cases n= 856 vs controls n=96,769). Carriers of large, rare (>500kb, <1% frequency) CNVs and known neurodevelopmental CNVs were identified. Primary analyses tested associations between early-onset depression and i) large, rare CNVs, and ii) neurodevelopmental CNVs. Secondary analyses investigated parent-reported measures of early-onset depression.

**Results:** Meta-analysis did not identify any robust associations between early-onset depression (SMFQ-defined) and large, rare CNVs (OR=0.92 [95% CI=0.84-1.02], p=0.12) or neurodevelopmental CNVs (OR=1.06 [0.85-1.31], p=0.60). No robust associations were observed between early-onset depression, defined using ICD-10 MDD criteria, and large rare CNVs (OR=1.08 [0.86-1.36], p=0.49) or neurodevelopmental CNVs (OR=0.69 [0.34-1.39], p=0.30).

**Conclusions:** Our findings did not support the hypothesis that individuals with early-onset depression show enrichment for large, rare or known neurodevelopmental CNVs.

## Introduction

Depression is a common psychiatric condition that is highly heterogeneous in its symptom presentation, age at onset, aetiology, clinical course, and treatment response (Goldberg, 2011). This substantial degree of heterogeneity has led to efforts to stratify depression into subtypes (Cai, Choi, & Fried, 2020). Several studies have suggested that an earlier age at onset of depression is one important indicator of phenotypic and genetic heterogeneity. Whilst the median age at onset of major depressive disorder is 30 years old, 13% of affected individuals experience depression onset by 18 years of age, and 3% by 14 years of age (Solmi et al., 2022). Depression that onsets in childhood or early adolescence is associated with a clinical course that is more chronic and recurrent, a greater likelihood of hospitalisation, and is more strongly associated with neurodevelopmental comorbidities such as autism spectrum diagnosis (ASD) and attention-deficit hyperactivity disorder (ADHD) than depression that onsets in later adolescence or adulthood (Dennison et al., 2024; Doering et al., 2022; Jaffee et al., 2002; van Os, Jones, Lewis, Wadsworth, & Murray, 1997).

Family and twin studies of depression suggest that age at onset indexes familial and genetic heterogeneity. The largest study to date, using Swedish registry data, found that youth-onset depression (<21 years) showed significantly higher heritability than adult-onset depression (>25 years) (Nguyen et al., 2023). Whilst genetic correlations between other depression subtypes (e.g. with vs without psychiatric comorbidity, impairment, suicidality) were substantial (rg=0.75-0.9), this was not the case for youth-onset vs adult-onset depression (rg=0.33), suggesting that youth-onset depression may represent a partially distinct depression subtype.

Molecular genetic studies offer the additional opportunity of providing biological insights into depression heterogeneity. Consistent evidence of distinct depression subtypes based on common genetic variant profiles (Howard et al., 2020) has not yet been found. However, higher polygenic scores for ADHD, bipolar disorder, and schizophrenia are associated with depression that onsets earlier in adolescence (Musliner et al., 2019; Weavers et al., 2021). These clinical and genetic studies have led to the hypothesis that child/early adolescent-onset depression may represent a “neurodevelopmental” subtype of depression (Rice et al., 2019).

Rare chromosomal micro-deletion and micro-duplication copy number variants (CNVs) are enriched in many neurodevelopmental conditions including intellectual disability/developmental delay, ASD, ADHD, and schizophrenia (Coe et al., 2014; Pinto et al., 2010; Rees et al., 2014; Williams et al., 2010). Evidence for association between known neurodevelopmental (ND) CNVs and depression however is inconsistent (Birnbaum, Mahjani, Loos, & Sharp, 2022; Kendall et al., 2019; Vaez et al., 2024). ND CNVs were associated with diagnosed depression in biobanks in the UK and the US (Birnbaum et al., 2022; Kendall et al., 2019), and with self-reported depression in the UK Biobank (Kendall et al., 2019). However, in the Danish iPSYCH sample, ND CNVs were not associated with diagnosed depression (Vaez et al., 2024). Given evidence of a higher neurodevelopmental burden, in terms of clinical phenotypes and common genetic variants, in very early-onset depression (childhood and early adolescence) and that an earlier age at onset indexes depression clinical and genetic heterogeneity, we aimed to further test the hypothesis that early-onset depression represents a “neurodevelopmental” depression subtype.

In this study we assembled data across four population-based cohorts from the UK, Norway, and Sweden, to examine whether large, rare CNVs and known ND CNVs are associated with early-onset depression.

## Methods

### Participants

Participants were ascertained from four population-based cohorts: 1) UK Avon Longitudinal Study of Parents and Children (ALSPAC), 2) UK Millennium Cohort Study (MCS), 3) Norwegian Mother, Father, and Child cohort Study (MoBa), and 4) Child and Adolescent Twin Study in Sweden (CATSS).

#### ALSPAC

Pregnant women residing in Avon, UK with an expected delivery date between 1^st^ April 1991 and 31^st^ December 1992 were invited to participate in ALSPAC. A total of 15,658 children were enrolled into the study across two waves of recruitment, of whom 14,901 were alive at 1 year of age (Boyd et al., 2013; Fraser et al., 2013; Northstone et al., 2019). The study website contains details of all the data that are available through a fully searchable data dictionary and variable search tool https://www.bristol.ac.uk/alspac/researchers/our-data/.

#### MCS

MCS recruited children born between 1st September 2000 and 11th January 2002 in sampling areas across the UK. Eligible children were identified through the child benefit register, which captured almost all children living in the UK at the time (Connelly & Platt, 2014). Initial recruitment was completed when children were 9 months old, with a second wave at 3 years resulting in a total sample size of 19,870 children.

#### MoBa

MoBa is a population-based pregnancy cohort study conducted by the Norwegian Institute of Public Health. Participants were recruited from all over Norway from 1999-2008. The women consented to participation in 41% of the pregnancies. The cohort includes approximately 114,500 children, 95,200 mothers and 75,200 fathers (Magnus et al., 2016).

#### CATSS

Since July 2004, all twins born in Sweden have been identified through the Swedish Twin Register and are contacted to participate in CATSS when they reach 9 years of age (Anckarsäter et al., 2011). Twins born between July 1992 and June 1995, i.e. those aged between 9 and 12 years at the start of recruitment, were contacted at age 12 to participate in the study.

### Ethical information

ALSPAC Ethics and Law Committee and the Local Research Ethics Committees provided ethical approval for the study. Informed consent for the use of data collected via questionnaires and clinics was obtained from participants following the recommendations of the ALSPAC Ethics and Law Committee at the time. Consent for biological samples has been collected in accordance with the Human Tissue Act (2004). Ethical approval for MCS was obtained from the NHS Research Ethics Committee and parents provided informed consent for their child to participate. MoBa is regulated by the Norwegian Health Registry Act. The establishment of MoBa and initial data collection was based on a license from the Norwegian Data Protection Agency and approval from The Regional Committees for Medical and Health Research Ethics. The MoBa cohort is currently regulated by the Norwegian Health Registry Act. The current study was approved by The Regional Committees for Medical and Health Research Ethics (2016/1226/REK sør-øst C). Ethical approval for CATSS was granted by the Karolinska Institute Ethical Review Board and the Regional Ethical Review Board in Stockholm. Parents provided written consent for their child to participate in CATSS.

### CNV Calling and Annotation

DNA was collected from blood samples provided between birth and 7 years in ALSPAC, and samples were genotyped using the Illumina HumanHap 500 quad array. In MCS, DNA samples were collected via saliva at age 14, and genotyped on the Illumina Global Screening Array-24 v1.0. DNA for MoBa participants was extracted from blood samples taken from the umbilical cord after birth (Paltiel et al., 2014). Samples were genotyped in batches across several arrays: Illumina HumanCoreExome 12v1.1 and 24v1.0, Global Screening Array MD v1.0 and v3.0, and InfiniumOmniExpress-24 v1.2 (Corfield et al., 2024). Twins recruited to CATSS from 2008 onwards provided saliva samples for DNA extraction; DNA was genotyped using the Illumina Infinium PsychArray-24 BeadChip.

CNVs were called in each individual cohort using PennCNV, as described previously (Dennison et al., 2025; Martin et al., 2019). CNV quality control was performed separately for each cohort, with the following inclusion filters applied: Log R Ratio (LRR) standard deviation (SD) <2.5 SD from mean, number of CNVs per individual <100, waviness factor <2.5 SD from mean, number of probes >=20, confidence score >=10. We examined large, rare CNVs (<1%) and known ND CNVs because both have been implicated as increasing risk for child neurodevelopmental conditions (Martin et al., 2019). For large, rare CNVs, plink v1.07 was used to identify CNVs >500kb with a frequency <1%. For ND CNVs, 54 CNVs (Coe et al., 2014) were identified using criteria described by Kendall et al. 2017 (Table S1). Most ND CNVs were required to span >50% of the critical region, defined for each CNV in Table S1, except for single gene CNVs where deletions must have covered at least one exon, whilst duplications had to cover the whole gene.

### Depression definition

Our primary outcome was early-onset depression (onset ≤14 years) defined using the self-reported Short Mood and Feelings Questionnaire (SMFQ). Participants scoring >11 on the self-reported SMFQ, the cut-point validated for depression in this age group (Angold et al., 1995; Thabrew, Stasiak, Bavin, Frampton, & Merry, 2018), were defined as having early adolescent-onset depression. The SMFQ was administered at the following ages: 10, 12, and 13 years in ALSPAC, 14 years in MCS and 14 years in MoBa. Self-reported SMFQ data were not collected in CATSS. As ALSPAC included multiple timepoints with eligible data, participants meeting the threshold at any time point were defined as having early-onset depression. We chose 14 years as the maximum age for early-onset in our study due to evidence that depression prevalence increases sharply after this age (Rice et al., 2019; Solmi et al., 2022), consistency with previous literature (Dennison et al., 2024; Weavers et al., 2021), and data availability across the cohorts.

Our secondary outcome was an episode of depression meeting ICD-10 criteria of major depressive disorder (MDD) with onset ≤14 years. In ALSPAC, ICD-10 depression was assessed using the Development and Wellbeing Assessment (DAWBA)(Goodman, Ford, Richards, Gatward, & Meltzer, 2000), a structured interview conducted with a parent to ascertain ICD-10 diagnoses. Participants meeting criteria for MDD at ages 10 or 13 were defined as having early-onset depression. MoBa and CATSS are linked to the Norwegian and Swedish patient registries, respectively, which report any ICD-10 diagnoses the person has received in a secondary care setting (e.g. hospital, outpatient clinics, or from contract specialists). Depression in children and adolescents is treated in secondary not primary care in these countries. Individuals receiving a diagnosis of a depressive episode (F32, F33) ≤14 years were defined as having early adolescent-onset depression, individuals without a diagnosis, or with an MDD diagnosis at an older age, were included as controls. An ICD-10 diagnosis of depression was not available in MCS.

Given that previous studies have highlighted low agreement between parent and child ratings of depressive symptoms (Baumgartner et al., 2020), and that the predictive validity of parent and child ratings differs across childhood and adolescence (Lewis et al., 2012), we conducted sensitivity checks using parent-reported SMFQ to define early-onset depression. Parent-reported early adolescent-onset depression was defined as a score >11 on the parent-reported SMFQ at ages 10, 12, or 13, in ALSPAC, and ages 9 or 12 in CATSS. Parent-reported data were not available in MCS or MoBa for the full SMFQ.

### Data availability

ALSPAC data are managed by the University of Bristol and are available to bona fide researchers upon application. Researchers can apply for ALSPAC data access on the University of Bristol website: https://www.bristol.ac.uk/alspac/researchers/access/. Phenotypic data for MCS are available to download freely from the UK Data Service https://doi.org/10.5255/UKDA-Series-2000031. To access genetic data for MCS, researchers must apply through the Centre for Longitudinal Studies https://cls.ucl.ac.uk/data-access-training/genetic-data-and-biological-samples/, where applications will be subject to ethical approval.

Data from MoBa and the Medical Birth Registry of Norway used in this study are managed by the Norwegian Institute of Public Health, and can be made available to researchers upon approval from the Regional Committees for Medical and Health Research Ethics (REC), compliance with the EU General Data Protection Regulation (GDPR) and approval from the data owners. The consent given by the participants does not allow for storage of data on an individual level in repositories or journals.

Researchers who want access to data sets for replication should apply through helsedata.no. Access to data sets requires approval from The Regional Committee for Medical and Health Research Ethics in Norway and an agreement with MoBa.

Due to restrictions under the Swedish Public Access to Information and Secrecy Act, the data are not publicly available. Data access may be granted to researchers upon request and following a secrecy review. Interested researchers should contact Patrik K.E. Magnusson, Department of Medical Epidemiology and Biostatistics, Karolinska Institutet.

### Analysis

Primary analyses tested for association between i) large, rare CNVs and SMFQ-defined early-onset depression, and ii) ND CNVs and SMFQ-defined early-onset depression. For ALSPAC and MCS, we performed logistic regressions to test these associations. For MoBa, we performed conditional logistic regression, conditioning on family ID, to account for relatedness. All regression models were run separately for each cohort, then meta-analysed using a fixed-effects model, calculated using the function ‘metagen’ in the R package ‘meta’. A fixed-effects model was used as depression was defined using the same measure, the same informant, and within the same age range across the cohorts. We tested for association between early-onset depression and standard deviation of the Log R Ratio, B Allele Frequency drift, waviness, and genotyping array in each cohort. Where an association with depression was observed (p<0.05), we included the appropriate variable as a covariate in all regression analyses for that cohort.

For the secondary analysis, logistic regressions (ALSPAC) and conditional logistic regressions (MoBa, CATSS) were used as above to test association between large rare CNVs and early-onset ICD-10 depression diagnosis, and the association between ND CNVs and early-onset ICD-10 depression diagnosis. Results across cohorts were meta-analysed as with the primary analyses.

Sensitivity analyses were conducted for parent-reported early-onset depression using logistic regressions in ALSPAC and CATSS, with results meta-analysed as above. Sex differences were examined by repeating the primary analyses in males-only and females-only subsets of the samples and comparing odds ratios and confidence intervals.

## Results

### Descriptives

A total of 107,930 individuals across all cohorts had genomic data that passed QC (ALSPAC n=8,360; MCS n=7,827; MoBa n=81,125; CATSS n=10,618). Of those passing QC, 32,382 individuals had self-reported SMFQ data and 97,625 had early-onset MDD diagnosis data.

13.5% of ALSPAC participants,15.9% of MCS participants and 21.2% of MoBa participants experienced self-reported early-onset depression, defined using the SMFQ. Rates of early-onset ICD-10 depression diagnosis, defined via healthcare records or structured interview, were 1.2% in ALSPAC, 0.8% in MoBa, and 0.9% in CATSS.

9.3% (range 7.5% - 10.7%) of individuals across all cohorts carried a large, rare CNV, whilst 1.6% (range 1.4 - 2.3%) of individuals carried an ND CNV. The number of individuals carrying CNVs with and without self-reported early-onset depression and early-onset MDD are presented in Table 1.

**Table 1.** Numbers of individuals in each cohort with early-onset depression (self-reported SMFQ) and early-onset MDD (clinical diagnosis or structured interview) with and without a large, rare CNV or an ND CNV. Cell counts <5 are not specified to maintain anonymity.

| Cohort | Depression assessment | Case-control status | Large, rare CNV |  | ND CNV |  |
| --- | --- | --- | --- | --- | --- | --- |
|  |  |  | Non-carrier | Carrier | Non-carrier | Carrier |
| ALSPAC | Self-reported depression | No depression | 4802 (91.6%) | 441 (8.4%) | 5126 (97.8%) | 117 (2.2%) |
|  |  | Depression | 747 (91.3%) | 71 (8.7%) | 796 (97.3%) | 22 (2.7%) |
|  | MDD diagnosis | No MDD | 5366 (91.3%) | 510 (8.7%) | 5744 (97.8%) | 132 (2.2%) |
|  |  | MDD | 66 (90.4%) | 7 (9.6%) | 73 (100%) | 0 (0%) |
| MoBa | Self-reported depression | No depression | 13,333 (90.8%) | 1353 (9.2%) | 14,485 (98.6%) | 201 (1.4%) |
|  |  | Depression | 3608 (91.1%) | 351 (8.9%) | 3904 (98.6%) | 55 (1.4%) |
|  | MDD diagnosis | No MDD | 72,733 (90.5%) | 7650 (9.5%) | 79,221 (98.6%) | 1162 (1.4%) |
|  |  | MDD | 617 (89.6%) | 72 (10.4%) | 682 (99.0%) | 7 (1.0%) |
| MCS | Self-reported depression | No depression | 5746 (89.0%) | 710 (11.0%) | 6322 (97.9%) | 137 (2.1%) |
|  |  | Depression | 1109 (91.1%) | 108 (8.9%) | 1190 (97.8%) | 27 (2.2%) |
| CATSS | MDD diagnosis | No MDD | 9718 (92.5%) | 792 (7.5%) | 10,338 (98.4%) | 172 (1.6%) |
|  |  | MDD | 88 (93.6%) | 6 (6.4%) | >89 (>94.7%) | <5 (<5.3%) |

### Early-onset depression – self-reported questionnaire

Large, rare CNV carrier status was not associated with self-reported early-onset depression, defined using the SMFQ, in the meta-analysis (OR=0.92 [95% CI=0.84-1.02], p=0.12) or in any individual cohort, with the exception of MCS, where carrying a large rare CNV was associated with reduced likelihood of early-onset depression (Table 2, Figure 1).

**Figure 1.**
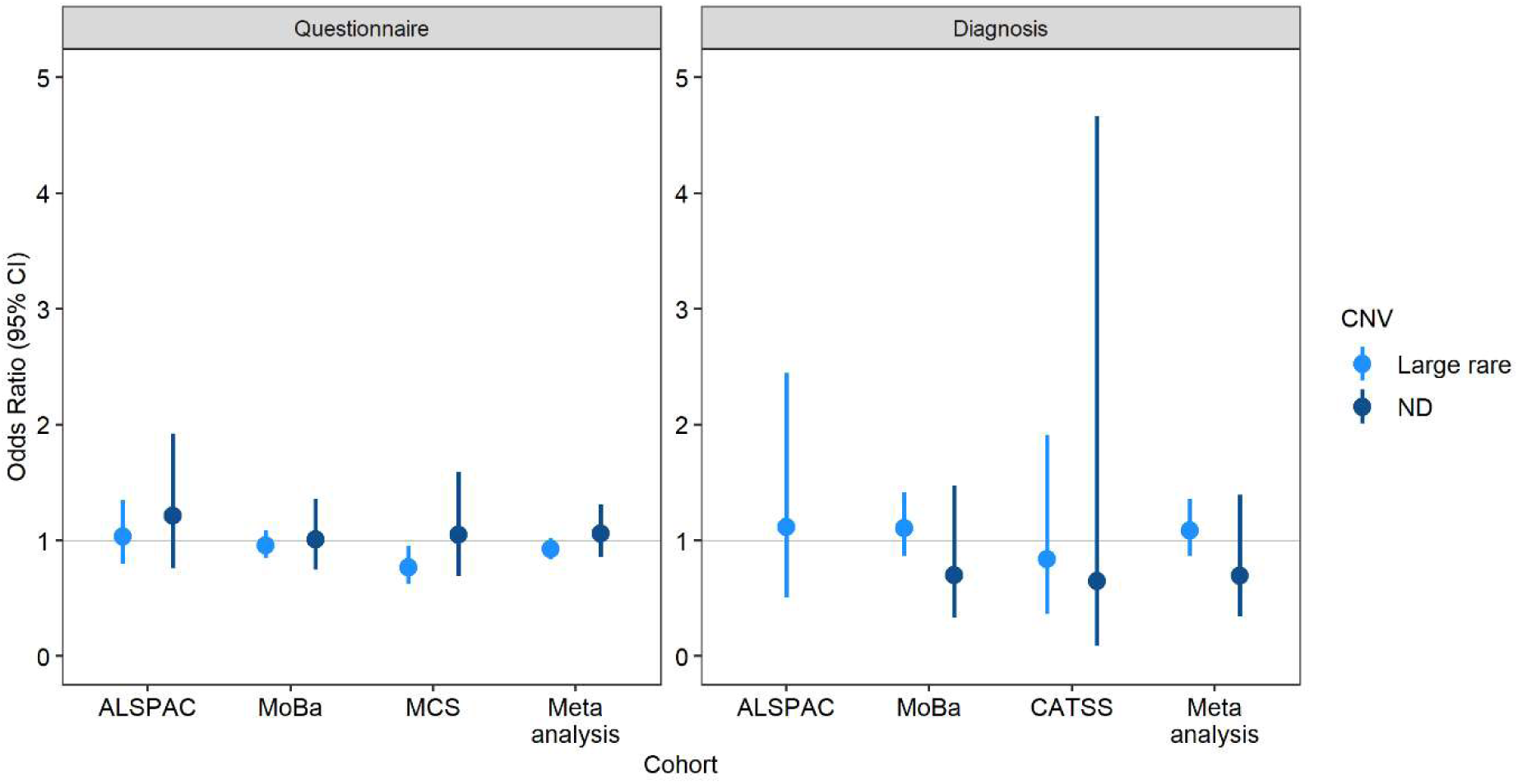
Results of analysis testing associations between early-onset depression and large rare CNVs (light blue) and neurodevelopmental (ND) CNVs (dark blue). Results are presented with a definition of depression based on either self-rated questionnaire (left panel) or as ICD-10 diagnosis (right panel). Points display Odds Ratio and bars refer to 95% confidence intervals.

**Table 2.** Association between CNVs and i) self-reported early-onset depression, defined via the SMFQ, and ii) early-onset MDD diagnosis.

| Analysis | Cohort | Large CNV |  |  |  | ND CNV |  |  |  |
| --- | --- | --- | --- | --- | --- | --- | --- | --- | --- |
|  |  | OR | Lower CI | Upper CI | P-value | OR | Lower CI | Upper CI | P-value |
| SMFQ self-reported depression | ALSPAC | 1.03 | 0.80 | 1.35 | 0.80 | 1.21 | 0.76 | 1.92 | 0.42 |
|  | MoBa | 0.96 | 0.85 | 1.08 | 0.50 | 1.01 | 0.75 | 1.36 | 0.97 |
|  | MCS | 0.77 | 0.62 | 0.95 | 0.01 | 1.05 | 0.69 | 1.59 | 0.83 |
|  | Meta-analysis | 0.92 | 0.84 | 1.02 | 0.12 | 1.06 | 0.85 | 1.31 | 0.60 |
| MDD diagnosis | ALSPAC | 1.12 | 0.51 | 2.45 | 0.78 | NA – insufficient cell count |  |  |  |
|  | MoBa | 1.10 | 0.86 | 1.41 | 0.43 | 0.70 | 0.33 | 1.48 | 0.35 |
|  | CATSS | 0.84 | 0.37 | 1.91 | 0.67 | 0.65 | 0.09 | 4.67 | 0.67 |
|  | Meta-analysis | 1.08 | 0.86 | 1.36 | 0.49 | 0.69 | 0.34 | 1.39 | 0.30 |

ND CNVs were not associated with depression in the meta-analysis (OR=1.06 [0.85-1.31], p=0.60), or in any individual cohort (Table 2, Figure 1).

### Early-onset MDD diagnosis

Large rare CNVs were not associated with early-onset depression meeting ICD-10 criteria for MDD in any cohort or the meta-analysis (OR=1.08 [0.86-1.36], p=0.49). No association was observed between ND CNVs and early-onset depression defined by ICD-10 MDD criteria in the meta-analysis (OR=0.69 [0.34-1.39], p=0.30) or any individual cohort (Table 2, Figure 1).

### Sensitivity analyses – parent-rated, SMFQ-defined depression

No evidence of association was observed between parent-reported early-onset depression and large, rare CNVs in ALSPAC or CATSS, or when the two cohorts were meta-analysed (Figure 2 and Table S2).

**Figure 2.**
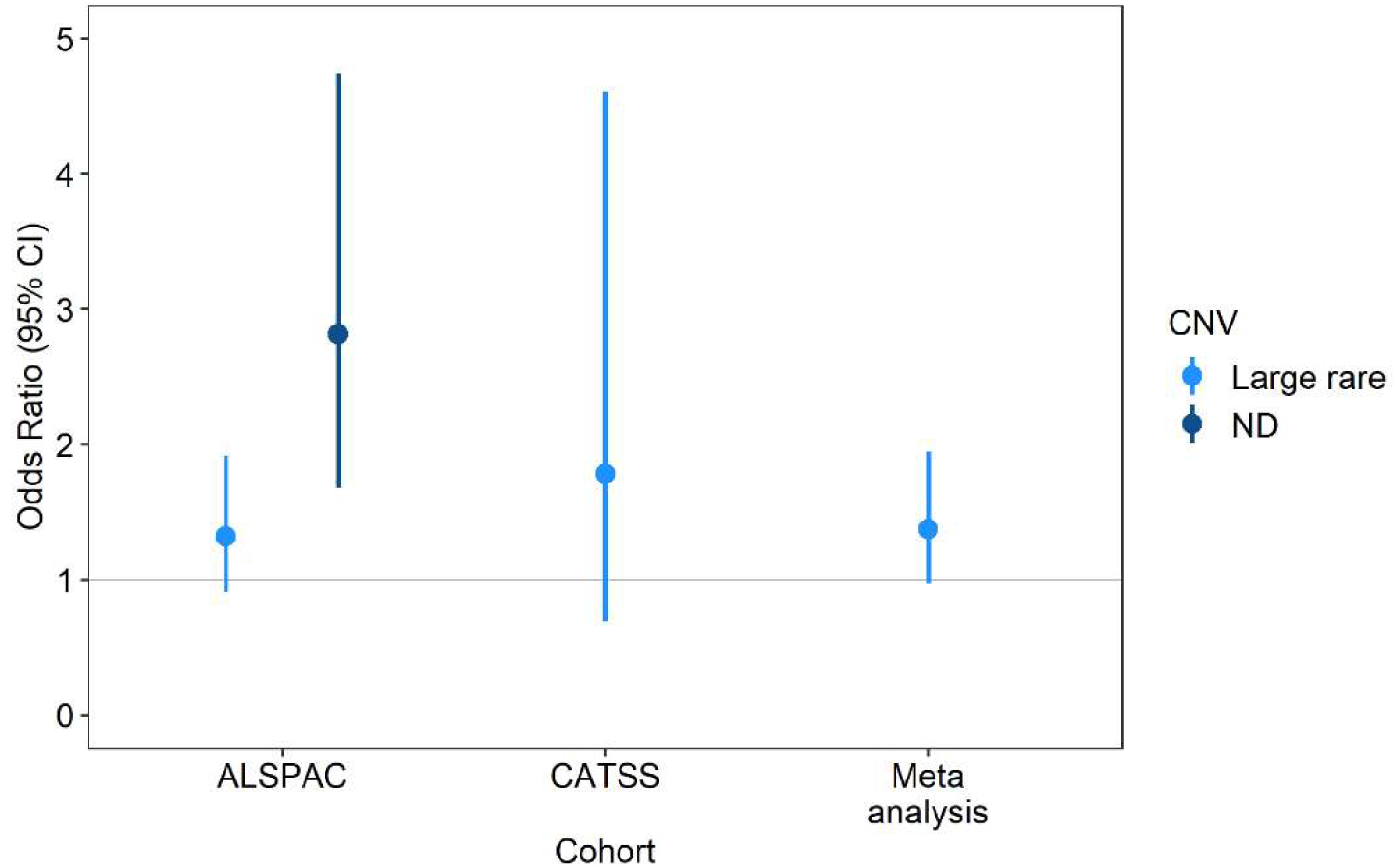
Results of analysis testing associations between parent-reported, questionnaire-defined early-onset depression and large rare (light blue) or neurodevelopmental (ND, dark blue) CNVs. Points display Odds Ratio and bars refer to 95% confidence intervals.

Parent-reported early-onset depression was associated with carrying an ND CNV in ALSPAC, but there were insufficient data to test this association in CATSS thus a meta-analysis could not be performed (Figure 2 and Table S2).

### Sex differences

Full results of the analyses examining sex differences are in Tables S3-S4 and Figure S1. There was evidence that females carrying a large rare CNV had a reduced likelihood of self-reported early-onset depression, which was not observed in males although the confidence intervals for males and females overlapped (Table S3 and Figure S1). No other differences were observed between males and females.

## Discussion

Across four population cohorts totalling 107,930 children and adolescents, we did not find strong evidence to support the hypothesis that rare CNVs are associated with early-onset depression ≤14 years). No associations were observed between large rare or ND CNVs and depression whether defined by self-reported SMFQ, or as an episode of depression meeting criteria for an ICD-10 MDD diagnosis. Our findings suggest that rare CNVs do not play a substantial role in the aetiology of early adolescent-onset depression.

There is some evidence to suggest that ND CNVs may be associated with depression in adulthood, although findings are inconsistent (Birnbaum et al., 2022; Kendall et al., 2019; Vaez et al., 2024). Nevertheless, CNVs have been consistently associated with an elevated likelihood of neurodevelopmental conditions, including intellectual disability, ASD, and ADHD, that themselves are associated with earlier-onset depression (Dennison et al., 2024; Doering et al., 2022). There are several reasons that may explain why we did not find evidence of association between ND CNVs and early-onset depression.

First, the association between early-onset depression and neurodevelopmental conditions may be explained by shared common genetic liability rather than by rare copy number variants.

Developmental trajectories of self-reported depression symptoms across childhood, adolescence, and adulthood suggest that earlier onset, persistent depression is associated with elevated ADHD polygenic scores (Weavers et al., 2021). Mendelian randomisation studies also suggest a causal relationship between common genetic variants for ADHD and risk of depression (Riglin et al., 2021). Thus, this could suggest that the neurodevelopmental component to early-onset depression is mainly driven by common genetic variation, rather than rare genetic variation.

Another possibility is that the higher heritability estimates and worse outcomes associated with early-onset compared to later-onset depression may be explained by early environmental adversities that are correlated with genetic risk (i.e. gene-environment correlation). For instance, adverse childhood experiences, which are increased for children with neurodevelopmental conditions (McDonnell et al., 2019), show correlation with genetic liability (Baldwin et al., 2023; Warrier et al., 2021), and are associated with depression risk (Hughes et al., 2017). Additionally, it is possible that depression may be missed or under-reported when the individual has complex needs or other urgent health concerns, as is common among CNV carriers who can experience a range of neurodevelopmental and physical health impacts (Chawner, Watson, & Owen, 2021). Evidence also suggests that questionnaires assessing depressive symptoms, which are typically validated in neurotypical populations, may not be equally appropriate or valid for neurodivergent individuals (Cassidy, Bradley, Bowen, Wigham, & Rodgers, 2018), who are more likely to carry a CNV.

Our study relied primarily on self-reported depression data. Research typically suggests combining both parent and self-reports for most reliable assessment of youth depression (Cohen, So, Young, Hankin, & Lee, 2019). However, in longitudinal cohorts it is rare to have both parent and self-reports of the same measure assessed at the same time. The age range of participants included in our study covers a period where clinicians and healthcare professionals may move from parent-report to self-report (Youngstrom et al., 2011), and it is possible that the point at which self-report becomes a more reliable informant differs for different individuals and especially for neurodivergent individuals. In particular, adolescents with ADHD have been found to under-report depressive symptoms compared to parent reports, whilst neurotypical adolescents were found to over-report depressive symptoms compared to parent reports (Fraser et al., 2018). Indeed, we found evidence in the ALSPAC cohort to suggest that ND CNVs may be associated with parent-reported depression. However, we were unable to investigate this in the other samples due to lack of data availability. It is important for future studies to consider the impact of informant and measurement during the study design, and particularly how differences across ages and cohorts may affect the findings.

A strength of our study was the consistent measurement of depression across multiple cohorts – i.e. the same informant and the same questionnaire within the same age range, which is often difficult to achieve across population cohorts. Despite the large overall sample size, small individual numbers in specific cohorts prohibited some analyses, such as MDD diagnosis and ND CNVs in ALSPAC. Rare CNVs are uncommon by definition and depression onsets at or before 14 years is also rare. Thus, we cannot rule out the possibility that our null finding is a type I error explained by insufficient statistical power. However, power analysis indicated that we had 80% power to detect an odds ratio of 1.27 or larger for the association between ND CNVs and SMFQ-defined early-onset depression. Whilst effect sizes smaller than this could be statistically significant in a larger sample, they would not be meaningful enough to justify recommending routine genetic testing. Due to the rare nature of ND CNVs, we were unable to measure associations with specific types of CNVs and instead created a pooled measure of CNVs known to be associated with neurodevelopmental conditions (Coe et al., 2014). It is possible that some types of CNVs that we were unable to analyse (e.g. CNVs in specific loci, rarer CNVs), may be associated with early-onset depression.

Overall, our findings suggest that rare CNVs do not substantially increase the likelihood of early-onset depression, when assessed via self-report or ICD-10 diagnosis. Our findings are inconsistent with evidence of enrichment of neurodevelopmental phenotypes in early-onset depression (Dennison et al., 2024; Doering et al., 2022), but are consistent with some, although not all, evidence from CNV studies of depression in adult populations (Vaez et al., 2024).

## Acknowledgements and funding

This publication is the work of the authors and CD and AT will serve as guarantors for the contents of this paper. For the purpose of open access, the authors have applied a CC BY public copyright license to any Author Accepted Manuscript version arising.

This work was supported by the Wolfson Centre for Young People’s Mental Health, established with support from the Wolfson Foundation. JM is funded by the Welsh Government through Health and Care Research Wales via an NIHR Advanced Fellowship (NIHR-FS(A)-2022). IES is supported by South-Eastern Norway Regional Health Authority (#2020060), and Kristian Gerhard Jebsen Stiftelsen (SKGJ-MED-021) as well as the Norwegian ADHD research network. AH is supported by the Research Council of Norway (#336085) and the South-Eastern Norway Regional Health Authority (#2020020, #2020024).

The Norwegian Mother, Father and Child Cohort Study is supported by the Norwegian Ministry of Health and Care Services and the Ministry of Education and Research. We are grateful to all the participating families in Norway who take part in this on-going cohort study. We thank the Norwegian Institute of Public Health (NIPH) for generating high-quality genomic data. This research is part of the HARVEST collaboration, supported by the Research Council of Norway (#229624). We also thank the NORMENT Centre for providing genotype data, funded by the Research Council of Norway (#223273), Southeast Norway Health Authorities and Stiftelsen Kristian Gerhard Jebsen. We further thank the Centre for Diabetes Research, the University of Bergen for providing genotype data and performing quality control and imputation of the data funded by the ERC AdG project SELECTionPREDISPOSED, Stiftelsen Kristian Gerhard Jebsen, Trond Mohn Foundation, the Research Council of Norway, the Novo Nordisk Foundation, the University of Bergen, and the Western Norway Health Authorities.

The analyses were also performed on resources on Services for sensitive data (TSD), University of Oslo, Norway, with resources provided by Sigma2 - the National Infrastructure for High-Performance Computing and Data Storage in Norway (projects NS9703S). Data from the Norwegian Patient Registry has been used in this publication. The interpretation and reporting of these data are the sole responsibility of the authors, and no endorsement by the Norwegian Patient Registry is intended nor should be inferred.

The UK Medical Research Council and Wellcome (Grant ref: 217065/Z/19/Z) and the University of Bristol provide core support for ALSPAC. For ALSPAC, genome-wide genotyping data was generated by Sample Logistics and Genotyping Facilities at Wellcome Sanger Institute and LabCorp (Laboratory Corporation of America) using support from 23andMe. A comprehensive list of grants funding is available on the ALSPAC website (http://www.bristol.ac.uk/alspac/external/documents/grant-acknowledgements.pdf). We are extremely grateful to all the families who took part in this study, the midwives for their help in recruiting them, and the whole ALSPAC team, which includes interviewers, computer and laboratory technicians, clerical workers, research scientists, volunteers, managers, receptionists and nurses.

We thank the Swedish Twin Registry for access to data. The Swedish Twin Registry is managed by Karolinska Institutet and receives funding through the Swedish Research Council (Grant No. 2017-00641).

## Correspondence to

Dr Charlotte Dennison

Hadyn Ellis Building, Maindy Road, Cardiff, CF24 4HQ, UK **Error! Hyperlink reference not valid.**

## References

Anckarsäter, H., Lundström, S., Kollberg, L., Kerekes, N., Palm, C., Carlström, E., Långström, N., et al. (2011). The Child and Adolescent Twin Study in Sweden (CATSS). Twin Research and Human Genetics, 14(6), 495–508.

Angold, A., Costello, E., Messer, S., Pickles, A., Winder, F., & Silver, D. (1995). Development of a short questionnaire for use in epidemiological studies of depression in children and adolescents. International Journal of Methods in Psychiatric Research, 5, 237–249.

Baldwin, J. R., Sallis, H. M., Schoeler, T., Taylor, M. J., Kwong, A. S. F., Tielbeek, J. J., Barkhuizen, W., et al. (2023). A genetically informed Registered Report on adverse childhood experiences and mental health. Nature Human Behaviour, 7(2), 269–290.

Baumgartner, N., Häberling, I., Emery, S., Strumberger, M., Nalani, K., Erb, S., Bachmann, S., et al. (2020). When parents and children disagree: Informant discrepancies in reports of depressive symptoms in clinical interviews. Journal of Affective Disorders, 272, 223–230.

Birnbaum, R., Mahjani, B., Loos, R. J. F., & Sharp, A. J. (2022). Clinical Characterization of Copy Number Variants Associated With Neurodevelopmental Disorders in a Large-scale Multiancestry Biobank. JAMA Psychiatry, 79(3), 250–259.

Boyd, A., Golding, J., Macleod, J., Lawlor, D. A., Fraser, A., Henderson, J., Molloy, L., et al. (2013). Cohort Profile: The ‘Children of the 90s’—the index offspring of the Avon Longitudinal Study of Parents and Children. International Journal of Epidemiology, 42(1), 111–127.

Cai, N., Choi, K. W., & Fried, E. I. (2020). Reviewing the genetics of heterogeneity in depression: Operationalizations, manifestations and etiologies. Human Molecular Genetics, 29(R1), R10– R18.

Cassidy, S. A., Bradley, L., Bowen, E., Wigham, S., & Rodgers, J. (2018). Measurement properties of tools used to assess depression in adults with and without autism spectrum conditions: A systematic review. Autism Research, 11(5), 738–754.

Chawner, S., Watson, C. J., & Owen, M. J. (2021). Clinical evaluation of patients with a neuropsychiatric risk copy number variant. Current Opinion in Genetics & Development, Molecular and genetic basis of disease, 68, 26–34.

Coe, B. P., Witherspoon, K., Rosenfeld, J. A., van Bon, B. W. M., Vulto-van Silfhout, A. T., Bosco, P., Friend, K. L., et al. (2014). Refining analyses of copy number variation identifies specific genes associated with developmental delay. Nature Genetics, 46(10), 1063–1071. Nature Publishing Group.

Cohen, J. R., So, F. K., Young, J. F., Hankin, B. L., & Lee, B. A. (2019). Youth Depression Screening with Parent and Self-Reports: Assessing Current and Prospective Depression Risk. Child psychiatry and human development, 50(4), 647–660.

Connelly, R., & Platt, L. (2014). Cohort Profile: UK Millennium Cohort Study (MCS). International Journal of Epidemiology, 43(6), 1719–1725.

Corfield, E. C., Shadrin, A. A., Frei, O., Rahman, Z., Lin, A., Athanasiu, L., Akdeniz, B. C., et al. (2024, May 7). The Norwegian Mother, Father, and Child cohort study (MoBa) genotyping data resource: MoBaPsychGen pipeline v.1. bioRxiv. Retrieved July 2, 2024, from https://www.biorxiv.org/content/10.1101/2022.06.23.496289v4

Dennison, C. A., Martin, J., Shakeshaft, A., Riglin, L., Powell, V., Kirov, G., Owen, M. J., et al. (2025). Early manifestations of neurodevelopmental copy number variants in children: A population-based investigation. Biological Psychiatry. Retrieved April 16, 2025, from https://www.sciencedirect.com/science/article/pii/S0006322325010509

Dennison, C. A., Shakeshaft, A., Eyre, O., Tilling, K., Rice, F., & Thapar, A. (2024). Investigating the neurodevelopmental correlates of early adolescent-onset emotional problems. Journal of Affective Disorders, 364, 212–220.

Doering, S., Halldner, L., Larsson, H., Gillberg, C., Kuja-Halkola, R., Lichtenstein, P., & Lundström, S. (2022). Childhood-onset versus adolescent-onset anxiety and depression: Epidemiological and neurodevelopmental aspects. Psychiatry Research, 312, 114556.

Fraser, A., Cooper, M., Agha, S. S., Collishaw, S., Rice, F., Thapar, A., & Eyre, O. (2018). The presentation of depression symptoms in attention-deficit/hyperactivity disorder: Comparing child and parent reports. Child and Adolescent Mental Health, 23(3), 243–250.

Fraser, A., Macdonald-Wallis, C., Tilling, K., Boyd, A., Golding, J., Davey Smith, G., Henderson, J., et al. (2013). Cohort Profile: The Avon Longitudinal Study of Parents and Children: ALSPAC mothers cohort. International Journal of Epidemiology, 42(1), 97–110.

Goldberg, D. (2011). The heterogeneity of “major depression”. World Psychiatry, 10(3), 226–228.

Goodman, R., Ford, T., Richards, H., Gatward, R., & Meltzer, H. (2000). The Development and Well-Being Assessment: Description and Initial Validation of an Integrated Assessment of Child and Adolescent Psychopathology. Journal of Child Psychology and Psychiatry, 41(5), 645–655.

Howard, D. M., Folkersen, L., Coleman, J. R. I., Adams, M. J., Glanville, K., Werge, T., Hagenaars, S. P., et al. (2020). Genetic stratification of depression in UK Biobank. Translational Psychiatry, 10(1), 163. Springer Nature.

Hughes, K., Bellis, M. A., Hardcastle, K. A., Sethi, D., Butchart, A., Mikton, C., Jones, L., et al. (2017). The effect of multiple adverse childhood experiences on health: A systematic review and meta-analysis. The Lancet Public Health, 2(8), e356–e366. Elsevier.

Jaffee, S. R., Moffitt, T. E., Caspi, A., Fombonne, E., Poulton, R., & Martin, J. (2002). Differences in Early Childhood Risk Factors for Juvenile-Onset and Adult-Onset Depression. Archives of General Psychiatry, 59(3), 215–222.

Kendall, K., Rees, E., Bracher-Smith, M., Legge, S., Riglin, L., Zammit, S., O’Donovan, M. C., et al. (2019). Association of Rare Copy Number Variants With Risk of Depression. JAMA Psychiatry, 76(8), 818–825.

Lewis, K. J. S., Mars, B., Lewis, G., Rice, F., Sellers, R., Thapar, A. K., Craddock, N., et al. (2012). Do parents know best? Parent-reported vs. child-reported depression symptoms as predictors of future child mood disorder in a high-risk sample. Journal of Affective Disorders, 141(2), 233–236.

Magnus, P., Birke, C., Vejrup, K., Haugan, A., Alsaker, E., Daltveit, A. K., Handal, M., et al. (2016). Cohort Profile Update: The Norwegian Mother and Child Cohort Study (MoBa). International Journal of Epidemiology, 45(2), 382–388.

Martin, J., Tammimies, K., Karlsson, R., Lu, Y., Larsson, H., Lichtenstein, P., & Magnusson, P. K. E. (2019). Copy number variation and neuropsychiatric problems in females and males in the general population. American Journal of Medical Genetics Part B: Neuropsychiatric Genetics, 180(6), 341–350.

McDonnell, C. G., Boan, A. D., Bradley, C. C., Seay, K. D., Charles, J. M., & Carpenter, L. A. (2019). Child maltreatment in autism spectrum disorder and intellectual disability: Results from a population-based sample. Journal of Child Psychology and Psychiatry, 60(5), 576–584.

Musliner, K. L., Mortensen, P. B., McGrath, J. J., Suppli, N. P., Hougaard, D. M., Bybjerg-Grauholm, J., Bækvad-Hansen, M., et al. (2019). Association of Polygenic Liabilities for Major Depression, Bipolar Disorder, and Schizophrenia With Risk for Depression in the Danish Population. JAMA Psychiatry, 76(5), 516–525.

Nguyen, T.-D., Kowalec, K., Pasman, J., Larsson, H., Lichtenstein, P., Dalman, C., Sullivan, P. F., et al. (2023). Genetic Contribution to the Heterogeneity of Major Depressive Disorder: Evidence From a Sibling-Based Design Using Swedish National Registers. The American journal of psychiatry, 180(10), 714–722.

Northstone, K., Lewcock, M., Groom, A., Boyd, A., Macleod, J., Timpson, N. J., & Wells, N. (2019). The Avon Longitudinal Study of Parents and Children (ALSPAC): An update on the enrolled sample of index children in 2019. Wellcome Open Research. Retrieved July 8, 2024, from https://wellcomeopenresearch.org/articles/4-51/v1

van Os, J., Jones, P., Lewis, G., Wadsworth, M., & Murray, R. (1997). Developmental Precursors of Affective Illness in a General Population Birth Cohort. Archives of General Psychiatry, 54(7), 625–631.

Paltiel, L., Haugan, A., Skjerden, T., Harbak, K., Bækken, S., Stensrud, N., Knudsen, G. P., et al. (2014). The biobank of the Norwegian Mother and Child Cohort Study—Present status. Norsk Epidemiologi, 24(1–2), 29–35.

Pinto, D., Pagnamenta, A. T., Klei, L., Anney, R., Merico, D., Regan, R., Conroy, J., et al. (2010). Functional impact of global rare copy number variation in autism spectrum disorders. Nature, 466(7304), 368–372. Nature Publishing Group.

Rees, E., Walters, J. T. R., Georgieva, L., Isles, A. R., Chambert, K. D., Richards, A. L., Mahoney-Davies, G., et al. (2014). Analysis of copy number variations at 15 schizophrenia-associated loci. The British Journal of Psychiatry, 204(2), 108–114. Cambridge University Press.

Rice, F., Riglin, L., Thapar, A. K., Heron, J., Anney, R., O’Donovan, M. C., & Thapar, A. (2019). Characterizing Developmental Trajectories and the Role of Neuropsychiatric Genetic Risk Variants in Early-Onset Depression. JAMA Psychiatry, 76(3), 306–313.

Riglin, L., Leppert, B., Dardani, C., Thapar, A. K., Rice, F., O’Donovan, M. C., Smith, G. D., et al. (2021). ADHD and depression: Investigating a causal explanation. Psychological Medicine, 51(11), 1890–1897. Cambridge University Press.

Solmi, M., Radua, J., Olivola, M., Croce, E., Soardo, L., Salazar de Pablo, G., Il Shin, J., et al. (2022). Age at onset of mental disorders worldwide: Large-scale meta-analysis of 192 epidemiological studies. Molecular Psychiatry, 27(1), 281–295. Nature Publishing Group.

Thabrew, H., Stasiak, K., Bavin, L.-M., Frampton, C., & Merry, S. (2018). Validation of the Mood and Feelings Questionnaire (MFQ) and Short Mood and Feelings Questionnaire (SMFQ) in New Zealand help-seeking adolescents. International Journal of Methods in Psychiatric Research, 27(3), e1610.

Vaez, M., Montalbano, S., Calle Sánchez, X., Georgii Hellberg, K.-L., Dehkordi, S. R., Krebs, M. D., Meijsen, J., et al. (2024). Population-Based Risk of Psychiatric Disorders Associated With Recurrent Copy Number Variants. JAMA Psychiatry. Retrieved July 4, 2024, from 10.1001/jamapsychiatry.2024.1453

Warrier, V., Kwong, A. S. F., Luo, M., Dalvie, S., Croft, J., Sallis, H. M., Baldwin, J., et al. (2021). Gene– environment correlations and causal effects of childhood maltreatment on physical and mental health: A genetically informed approach. The Lancet Psychiatry, 8(5), 373–386. Elsevier.

Weavers, B., Heron, J., Thapar, A. K., Stephens, A., Lennon, J., Bevan Jones, R., Eyre, O., et al. (2021). The antecedents and outcomes of persistent and remitting adolescent depressive symptom trajectories: A longitudinal, population-based English study. The Lancet. Psychiatry, 8(12), 1053–1061.

Williams, N. M., Zaharieva, I., Martin, A., Langley, K., Mantripragada, K., Fossdal, R., Stefansson, H., et al. (2010). Rare chromosomal deletions and duplications in attention-deficit hyperactivity disorder: A genome-wide analysis. The Lancet, 376(9750), 1401–1408. Elsevier.

Youngstrom, E. A., Youngstrom, J. K., Freeman, A. J., De Los Reyes, A., Feeny, N. C., & Findling, R. L. (2011). Informants Are Not All Equal: Predictors and Correlates of Clinician Judgments About Caregiver and Youth Credibility. Journal of Child and Adolescent Psychopharmacology, 21(5), 407–415. Mary Ann Liebert, Inc., publishers.

