## Supplementary material for "The role of rare copy number variants in early-onset depression": Table S1

**Table S1** CNV calling criteria

| CNV | Coordinates (hg19) | Criteria |
|---|---|---|
| 1p36 del (GABRD) | chr1:0-2500000 | Size >50% of critical region, affecting GABRD |
| 1p36 dup (GABRD) | chr1:0-2500000 | Size >50% of critical region, affecting GABRD |
| TAR del 1q21.2 | chr1:145,394,955-145,807,817 | Size >50% of critical region |
| TAR dup 1q21.1 | chr1:145,394,955-145,807,817 | Size >50% of critical region |
| 1q21.1 del | chr1:146,527,987-147,394,444 | Size >50% of critical region |
| 1q21.1 dup | chr1:146,527,987-147,394,444 | Size >50% of critical region |
| NRXN1 del 2p16.3 | chr2:50145643-51259674 | Exonic deletions |
| 2q11.2 del (LMAN2L, ARID5A) | chr2:96,742,409-97,677,516 | Size >50% of critical region, affecting both LMAN2L and ARID5A |
| 2q13 del | chr2:111,394,040-112,012,649 | Size >50% of critical region |
| 2q13 dup | chr2:111,394,040-112,012,649 | Size >50% of critical region |
| 2q37 del (HDAC4) | chr2:239,716,679-243,199,373 | Size >50% of critical region, affecting HDAC4 |
| 3q29 del | chr3:195,720,167-197,354,826 | Size >50% of critical region |
| Wolf-Hirschhorn del (4p16.3) | chr4:1,552,030-2,091,303 | Size >50% of critical region |
| Wolf-Hirschhorn dup (4p16.3) | chr4:1,552,030-2,091,303 | Size >50% of critical region |
| Sotos syndrome del (5q35) | chr5:175,720,924-177,052,594 | Size >50% of critical region |
| Williams-Beuren syndrome del (7q11.23) | chr7:72,744,915-74,142,892 | Size >50% of critical region |
| Williams-Beuren syndrome dup (7q11.23) | chr7:72,744,915-74,142,892 | Size >50% of critical region |
| 8p23.1 del | chr8:8,098,990-11,872,558 | At least 1Mbp of critical region |
| 8p23.1 dup | chr8:8,098,990-11,872,558 | At least 1Mbp of critical region |
| 9q34 del (EHMT1) | chr9:140,513,444-140,730,578 | At least 1Mbp CNVs, including EHMT1 |
| 10q23 del (NRG3, GRID1) | chr10:82,045,472-88,931,651 | At least 1Mbp, including NRG3 and GRID1 |
| Potocki-Shaffer syndrome del (EXT2) (11p11.2) | chr11:43,940,000-46,020,000 | Size >50% of critical region, including EXT2 |
| 15q11.2 del BP1-BP2 | chr15:22,805,313-23,094,530 | Size >50% of critical region PWS del/dup Full critical region, ~4Mbp |
| 15q11.2 dup BP1-BP2 | chr15:22,805,313-23,094,530 | Size >50% of critical region PWS del/dup Full critical region, ~4Mbp |
| Prader-Willi syndrome/Angelman syndrome del (15q11.2q12) | chr15:22,805,313-28390339 | Full critical region, ~4Mbp |
| Prader-Willi syndrome/Angelman syndrome dup (15q11.2q12) | chr15:22,805,313-28390339 | Full critical region, ~4Mbp |
| 15q13.3 del BP4-BP5 | chr15:31,080,645-32,462,776 | Size >50% of critical region |
| 15q24 del | chr15:72900171-78151253 | At least 1Mbp between the A-E intervals |
| 15q24 dup | chr15:72900171-78151253 | At least 1Mbp between the A-E intervals |

| | | |
|---|---|---|
| 15q25 del | chr15:83,219,735-85,722,039 | At least 1Mbp between the A-D intervals |
| 16p13.11 del | chr16:15,511,655-16,293,689 | Size >50% of critical region |
| 16p13.11 dup | chr16:15,511,655-16,293,689 | Size >50% of critical region |
| 16p12.1 del | chr16:21,950,135-22,431,889 | Size >50% of critical region |
| 16p11.2 distal del | chr16:28,823,196-29,046,783 | Size >50% of critical region |
| 16p11.2 distal dup | chr16:28,823,196-29,046,783 | Size >50% of critical region |
| 16p11.2 del | chr16:29,650,840-30,200,773 | Size >50% of critical region |
| 16p11.2 dup | chr16:29,650,840-30,200,773 | Size >50% of critical region |
| 17p13.3 del (YWHAE) | chr17:1,247,834-1,303,556 | Exonic deletions; whole gene duplications |
| 17p13.3 dup (YWHAE) | chr17:1,247,834-1,303,556 | Exonic deletions; whole gene duplications |
| 17p13.3 del (PAFAH1B1) | chr17:2,496,923-2,588,909 | Exonic deletions; whole gene duplications |
| 17p13.3 dup (PAFAH1B1) | chr17:2,496,923-2,588,909 | Exonic deletions; whole gene duplications |
| Potocki-Lupski syndrome dup (17p11.2) | chr17:16,812,771-20,211,017 | Size >50% of critical region |
| Smith-Magenis syndrome del (17p11.2) | chr17:16,812,771-20,211,017 | Size >50% of critical region |
| 17q11.2 del (NF1) | chr17:29,107,491-30,265,075 | Size >50% of critical region, affecting NF1 |
| 17q11.2 dup (NF1) | chr17:29,107,491-30,265,075 | Size >50% of critical region, affecting NF1 |
| Renal cysts and diabetes syndrome del (17q12) | chr17:34,815,904-36,217,432 | Size >50% of critical region |
| 17q12 dup | chr17:34,815,904-36,217,432 | Size >50% of critical region |
| 17q21.31 del | chr17:43,705,356-44,164,691 | Size >50% of critical region |
| 22q11.2 del | chr22:19,037,332-21,466,726 | Size >50% of critical region |
| 22q11.2 dup | chr22:19,037,332-21,466,726 | Size >50% of critical region |
| 22q11.2 distal del | chr22:21,920,127-23,653,646 | Size >50% of critical region |
| 22q11.2 distal dup | chr22:21,920,127-23,653,646 | Size >50% of critical region |
| SHANK3 del (22q13) | chr22:51113070-51171640 | At least 1Mbp CNVs, including SHANK3 |
| SHANK3 dup (22q13) | chr22:51113070-51171640 | At least 1Mbp CNVs, including SHANK3 |

**Table S2.** Association between parent-reported early-onset depression (SMFQ-defined) and i) large rare CNVS, and ii) neurodevelopmental (ND) CNVs.

| Cohort | Large rare CNV | | | | ND CNV | | | |
|---|---|---|---|---|---|---|---|---|
| | OR | Lower CI | Upper CI | P-value | OR | Lower CI | Upper CI | P-value |
| ALSPAC | 1.32 | 0.91 | 1.92 | 0.14 | 2.82 | 1.67 | 4.74 | 9.7E-05 |
| CATSS | 1.78 | 0.69 | 4.61 | 0.23 | NA – insufficient cell count | | | |
| Meta-analysis | 1.37 | 0.97 | 1.94 | 0.07 | NA – only one cohort | | | |

**Table S3** Association between self-reported early-onset depression and large rare CNVs in males and females

| Cohort | Males | | | | Females | | | |
|---|---|---|---|---|---|---|---|---|
| | OR | Lower CI | Upper CI | P-value | OR | Lower CI | Upper CI | P-value |
| ALSPAC | 1.51 | 0.99 | 2.32 | 0.058 | 0.84 | 0.60 | 1.17 | 0.30 |
| MoBa | 0.96 | 0.75 | 1.24 | 0.78 | 0.94 | 0.81 | 1.09 | 0.39 |
| MCS | 0.99 | 0.69 | 1.44 | 0.97 | 0.71 | 0.54 | 0.93 | 0.013 |
| Meta-analysis | 1.06 | 0.88 | 1.28 | 0.55 | 0.76 | 0.61 | 0.94 | 0.010 |

**Table S4** Association between self-reported early-onset depression and neurodevelopmental CNVs in males and females

| Cohort | Males | | | | Females | | | |
|---|---|---|---|---|---|---|---|---|
| | OR | Lower CI | Upper CI | P-value | OR | Lower CI | Upper CI | P-value |
| ALSPAC | 1.15 | 0.52 | 2.54 | 0.73 | 1.33 | 0.75 | 2.36 | 0.33 |
| MoBa | 0.88 | 0.46 | 1.70 | 0.71 | 1.01 | 0.71 | 1.44 | 0.97 |
| MCS | 1.74 | 0.89 | 3.43 | 0.11 | 0.71 | 0.41 | 1.25 | 0.24 |
| Meta-analysis | 1.21 | 0.80 | 1.81 | 0.36 | 0.99 | 0.76 | 1.29 | 0.94 |

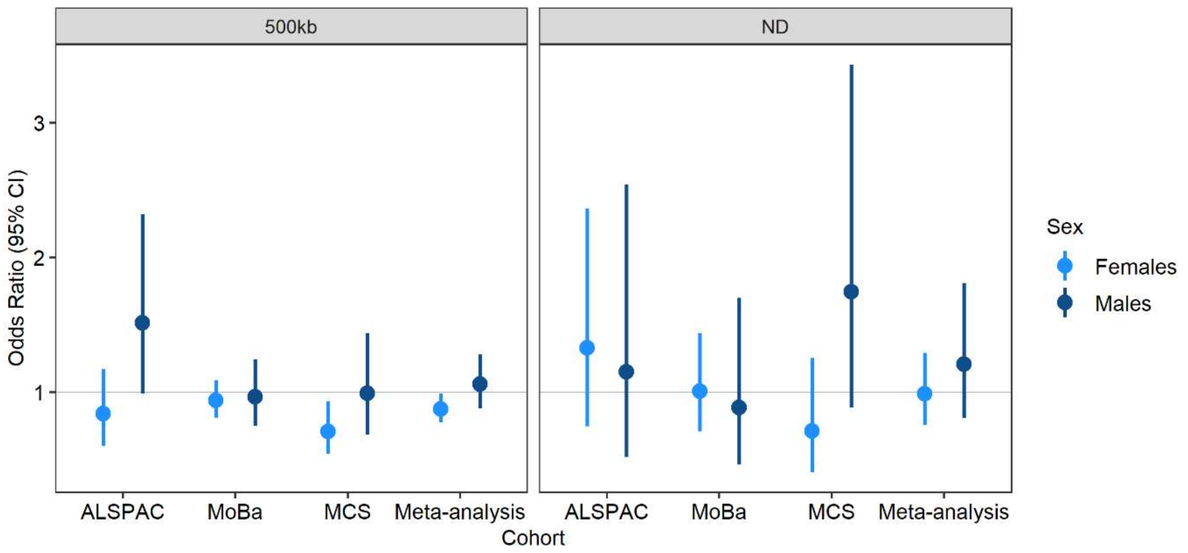

**Figure S1.** Association between self-reported early-onset depression and large rare CNVs (left panel) and neurodevelopmental CNVs (right panel) in females (light blue) and males (dark blue).
